# Exercise intensity shapes acute and prolonged immune and extracellular vesicle responses in older adults

**DOI:** 10.1101/2024.11.22.24317619

**Authors:** Alejandra P. Garza, Lorena Morton, Anna-Lena Motsch, Christian Puta, Marvin Stiebler, Yves Lading, Stefanie Schreiber, Rüdiger Braun-Dullaeus, Patrick Müller, Ildiko R. Dunay

**Affiliations:** Institute of Inflammation and Neurodegeneration, Otto-von-Guericke University, Germany; Department of Sports Medicine and Health Promotion, Friedrich-Schiller-University Jena, Jena, Germany; Department for Internal Medicine IV (Gastroenterology, Hepatology and Infectious Diseases), Jena University Hospital, Jena, Germany; Center for Sepsis Control and Care (CSCC), Jena University Hospital/Friedrich-Schiller-University Jena, Jena, Germany; Department of Cardiology and Angiology, University Hospital Magdeburg, Magdeburg, Germany; Center for Behavioral Brain Sciences (CBBS), Magdeburg, Germany; Department of Neurology, University Hospital Magdeburg, Magdeburg, Germany; German Center for Neurodegenerative Diseases (DZNE) Magdeburg; Department of Neurology, Heinrich Heine University Düsseldorf, Düsseldorf, Germany; German Center for Mental Health (DZPG), Magdeburg, Germany; Centre for Intervention and Research on Adaptive and Maladaptive Brain Circuits Underlying Mental Health (C-I-R-C), Magdeburg, Germany

**Keywords:** exercise, immunology, healthy aging, inflammaging, extracellular vesicles, exercise-induced immunomodulation, sex differences, aging and immune function, plasma biomarkers, cytokine response, personalized exercise

## Abstract

Regular physical activity is a cornerstone of healthy aging, offering a wide range of benefits, including the modulation of immune regulation and reduction of chronic inflammation. With aging closely linked to persistent, low-grade inflammation, i.e. inflammaging, the effects of exercise intensity on acute immune responses in older adults remain not fully understood. In this study, we explored how moderate and intense acute continuous exercise impact immune cell activation, cytokine production and large extracellular vesicle (lEV) release in healthy elderly individuals. Fourteen participants completed a moderate continuous exercise intervention (60% VO_2max_ for 30 minutes), while nineteen engaged in an intense continuous exercise session until exhaustion. Blood samples were collected at baseline, and at 1- and 24-hours post-exercise. Immune cell characterization by flow cytometry revealed distinct changes in monocyte subsets and NK cells activation across both exercise intensities. Intense exercise was associated with elevated proinflammatory TNFα levels, accumulation of circulating plasma-derived lEV and changes in their surface marker expression after 24 hours. Additionally, we identified sex-specific differences, including distinct activation profiles in innate immunity, alterations in EV release from CD4^+^ and HLA+ cells, and an exercise-induced increase in IL-6 observed exclusively in females. These findings suggest that moderate continuous acute exercise enhances immune cell activation without altering cell counts, while intense continuous exercise triggers acute proinflammatory immune response. Further research should clarify the long-term implications and fundamental mechanisms of exercise-induced immune modulation in aging populations.

**Key points summary:**

- Diminished immune function upon aging is increasing disease risk. This study examines how tailored acute exercise interventions stimulate immune regulation in older adults addressing age-related inflammatory challenges.
- Acute continuous moderate and intense exercise elicit distinct immune responses in elderly individuals with marked differences between sexes. Interestingly, IL-6 levels increased 30 min moderate exercise exclusively in females.
- Exercise promotes the release of extracellular vesicles (EVs) and modulates peripheral immunity, suggesting a potent mechanism by which physical activity supports immune resilience in aging.
- Tailored acute exercise regimens for older adults may enhance immune health, mitigating age-related inflammatory risks and enhancing resilience.
- This study emphasizes the need for further research on exercise-driven modulation focusing on sex differences and their implications for targeted interventions upon aging.

## Introduction

Aging is characterized by a progressive decline in physiological function and increased susceptibility to chronic diseases such as cardiovascular (e.g., heart failure, chronic coronary syndrome), metabolic (e.g., Diabetes Mellitus 2) and neurodegenerative (e.g., dementia) diseases, as well as cancer^1–5^. Age- related immune changes, or immunosenescence, contribute to heightened disease vulnerability and impaired infection response^6^. Chronic low-grade inflammation, or *inflammaging*, is a hallmark of aging and contributes to the pathogenesis of age-related diseases^7,8^. As the global population ages, understanding the complex interplay between exercise, immune function, inflammation, and aging is essential to promote healthy aging and reduce the burden of age-related diseases^9^.

Physical activity confers numerous health benefits and is essential for promoting healthy aging. While evidence supports that exercise positively impacts outcomes and prognoses across numerous diseases in the elderly^10^, the underlying mechanisms of exercise-induced release, uptake, and communication of bioactive factors in older adults remain incompletely understood. This is especially true regarding sex-specific responses as males and females may exhibit distinct immune adaptations to exercise. Regular exercise modulates immune function by reducing systemic inflammation markers and attenuates age-related increases in proinflammatory cytokine levels, making it a powerful approach for combating inflammaging and mitigate chronic disease risk in older adults^7,9,11–14^. Importantly, sex-specific differences in immune function suggest that personalized exercise regimens could optimize immune responses and overall health outcomes, highlighting the need for targeted approaches in exercise prescriptions for aging populations^15^.

Age-related immune changes include alterations in immune cell composition and function, dysregulation of cytokine signaling, and impaired immune surveillance^16–20^. While the benefits of physical exercise on immune function are well-documented, the precise mechanisms driving these effects, particularly in the elderly population, remain incompletely understood.

In recent years, extracellular vesicles (EVs), particularly those released in response to exercise (ExerVs), have emerged as a subject of significant interest due to their role in facilitating intercellular communication. By transporting proteins, lipids, and microRNAs to distant organs, EVs act as carriers of bioactive molecules potentially mediating exercise-induced benefits across the organism. Evidence suggests that exercise modulates EV release into circulation pointing towards a pathway through which physical activity may influence immune responses in aging populations^21–26^.

Despite growing recognition of the importance of physical exercise in healthy aging, major gaps remain in our understanding of the mechanisms underlying its effects on immune function, inflammation, and aging, along with sex-related discrepancies^7,13,27^. Thus, our research aimed to investigate these gaps by examining the effects of acute continuous moderate and intense exercise interventions on immune responses, cytokine levels, and the release of ExerVs release in older adults. Furthermore, we aimed to examine the importance of sex-specific responses, focusing on how tailored exercise interventions may promote immune resilience and health in aging populations.

## Methods

### Study population

A cohort of 33 healthy individuals (subproject 1: 14 participants, subproject 2: 19 participants), recruited through the University Hospital Magdeburg, participated in this study. The inclusion criteria were age > 55 years and the ability to move freely. Participants with a history of cardiovascular, endocrinological, neurological, neoplastic, or psychiatric disorders were excluded.

### Intervention

***Subproject 1:*** Fourteen healthy adults (median age: 68 years, 57 % female) underwent a moderate acute continuous exercise intervention on a bicycle ergometer (30 minutes, 60 % VO_2max_). Blood was collected before and 30 minutes after exercise intervention (Figure 1A). Individual cardiorespiratory fitness was prior assessed using cardiopulmonary exercise testing.

**Figure 1.**
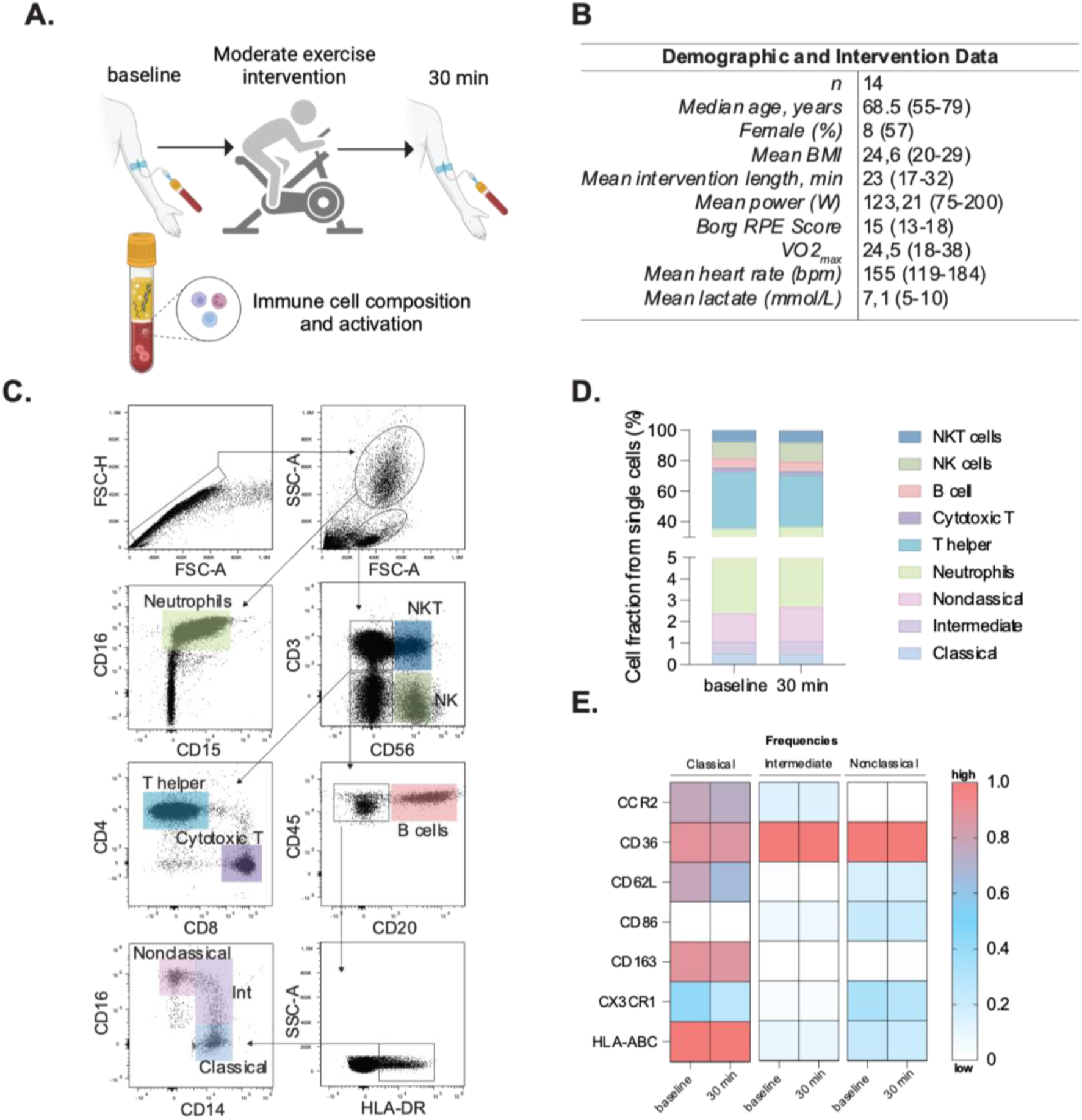
Study design and immune cell activation changes following acute continuous moderate exercise. Clinical and experimental study design and analytical processes for moderate exercise intervention. Peripheral blood was assessed at baseline and 30 min after the session **(A)**. Demographic and intervention data of all participants **(B)**. Representative gating strategy to identify single cells, followed by granulocytes and mononuclear cells based on size and granularity (SSC-A vs FSC-A). Neutrophils were gated based on FSC and SSC, followed by their expression of CD16 and CD15. Mononuclear cells were gated on the expression of CD3 and CD56, to obtain NKT cells (CD56^+^, CD3^+^), NK cells (CD56^+^, CD3^-^). From the double negative population (CD56^-^, CD3^-^), we assessed the presence of CD20 to represent B cells, and the CD20-cells, were further gated based on their surface expression of CD16 and CD14, where classical monocytes were defined as CD14^+^, CD16^-^; intermediate monocytes were CD14^+^, CD16^low^; and nonclassical monocytes as CD16^++^, CD14^low^ **(C)**. Bar charts showing cell fractions of all investigated immune cell populations at baseline and 30 min after the intervention **(D)**. Heatmaps showing the normalized frequency of key activation markers of monocyte subsets (CCR2, CD36, CD62L, CD86, CD163, CX3CR1 and HLA-ABC) **(E)**. Statistical evaluations using non-parametric student’s t-test. P values: * for *p* ≤ 0.05; ** for *p* ≤ 0.001; *** for *p* ≤ 0.0001. Panel A was created in BioRender. Garza, AP. (2024) https://BioRender.com/o96f904.

***Subproject 2:*** Nineteen healthy adults (median age: 67 years, 47 % female) performed a cardiopulmonary exercise test until exhaustion on a bicycle ergometer. The incremental step test included a three-minute unloaded pedaling at 0 W, following the resistance increased by 25 W every three minutes. During the incremental cycling test, breath-by-breath pulmonary gas-exchange data (MetaSoft, Studio: Cortex Biophysik GmbH Leipzig, Germany), heart rate (Custo med 100, custo med GmbH, Ottobrunn, Germany) and lactate levels (Lactate Scout 4, EKF Diagnostic, Barleben, Germany) were assessed. Perceived exertion was assessed at the end of each step using Borg-Scale (Borg, 1970). The cardiopulmonary exercise testing (CPET) concluded when (i) the respiratory exchange ratio was above 1.10, (ii) a plateau in VO2 occurred (despite increasing workload) or (iii) the rating of perceived exertion was 18 or higher on the Borg Scale. Safety criteria’s for premature termination of CPET were in the case of major electrocardiographic abnormalities, excessive blood pressure increase (≥ 230 mmHg systolic and/or ≥ 110 mmHg diastolic), or individual request^28^. Blood samples were collected at baseline, 30 min and 24 hours post-intervention. (Figure 2A).

**Figure 2.**
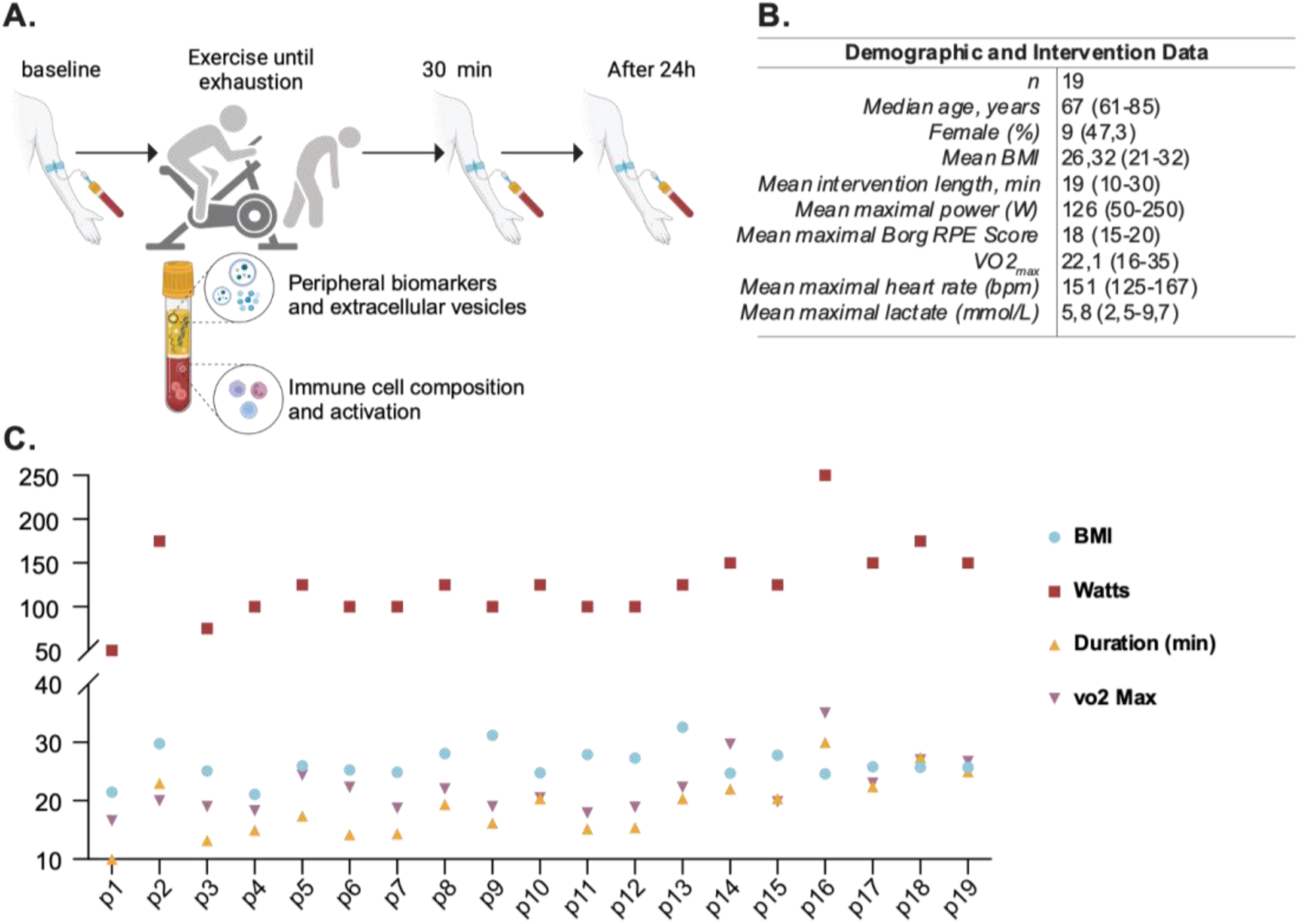
Study design and participant characteristics for intense exercise intervention. Clinical and experimental study design and analytical processes for moderate exercise intervention. Peripheral blood was assessed at baseline, 30 min, and 24 h after the intervention **(A)**. Demographic and intervention data of all participants **(B)**. Individual datapoints showing relevant variables in the cohort including body mass index (BMI), Watts reached, duration of the intervention in minutes, VO_2max_ and age **(C)**. Panel A was created in BioRender. Garza, AP. (2024) https://BioRender.com/c87a107.

### Blood collection and processing

Whole blood was obtained from the antecubital area via venipuncture using a 21G butterfly needle in sterile BD Vacutainer blood collection tubes containing 1 mL Acid Citrate Dextrose/Glucose (ACD). Samples were processed within 1 h of collection. Whole blood (100 µL) was lysed using 1X Red Blood Cell Lysis Buffer (BioLegend, 10X) following the manufacturer’s instructions. After lysis, cells were centrifuged at 400 × g for 5 min at room temperature. The supernatant was discarded, and two washing steps were performed using PBS. Cells were resuspended in 300 µL of FACS buffer (1X PBS, 2 % FBS, 2 mM EDTA, and 2 mM NaN3), and 100 µL was added to a 5 mL round-bottom polystyrene tube. Samples were incubated in 5 µL of Human TruStain FcX (BioLegend) for 10 min to avoid nonspecific antibody binding. Each sample was stained with the following antibodies: anti-human CD16 (FITC), anti-human HLA-DR (Peridinin chlorophyll protein-Cyanine5.5), anti-human CD86 (Allophycocyanin), anti-human CD3 (Alexa Fluor 700), anti-human CD66b (Alexa Fluor 700), anti-human CD19 (Alexa Fluor 700), anti-human CD56 (Alexa Fluor 700), anti-human CD36 (Allophycocyanin-Cyanine 7), anti-human CD163 (Brilliant Violet 421), anti-human CD15 (Brilliant Violet 510), anti-human HLA-ABC (Brilliant Violet 605), anti-human CCR2 (Phycoerythrin), anti-human CD62L (Phycoerythrin -Dazzle 594), anti-human CD15 (Phycoerythrin- Cyanine5), anti-human CX3CR1 (Phycoerythrin-Cy7), anti-human CD14 (Alexa Fluor 700), anti-human CD15 (APC), anti-human CD4 (Allophycocyanin-Cyanine 7), anti-human HLA-DR (Brilliant Violet 421), anti-human CD3 (Brilliant Violet 510), anti-human CD19 (Brilliant Violet 605), anti-human CD16 (Brilliant Violet711), anti-human CD45 (FITC), anti-human CD56 (Phycoerythrin), CD8 (Phycoerythrin-Dazzle 594) and anti-human CD20 (Peridinin chlorophyll protein-Cyanine5.5). After an incubation period of 30 min, the samples were washed twice and resuspended in 210 µL of FACS buffer. Samples were acquired using an Attune NxT flow cytometer (Thermo Fisher Scientific) and further analyzed using Flowjo (v10.10.0).

### Cytokine assay

Plasma was separated from whole blood by centrifugation at 1,500 g for 10 min. Plasma (1 mL) was transferred to clean Eppendorf tubes and centrifuged at 400 × g for 10 min at 4 °C to remove debris. Analytes were assessed using the Human LEGENDplex Multiplex Assay (BioLegend) following the manufacturer’s instructions. The assay utilizes allophycocyanin-coated beads conjugated with surface antibodies that allow the specific binding of the analyte of interest. After incubation of the capture beads with the plasma sample, biotinylated detection antibodies were added to create a bead-analyte-detection antibody sandwich, which was further stained with streptavidin-phycoerythrin. Samples were measured using an Attune NxT flow cytometer and analyzed using the LEGENDplex Data Analysis Online Software Suit. Half of the limit of detection (LOD) was used as a constant value when the predicted concentrations were below the LOD.

### Extracellular vesicles

Plasma samples from the intense exercise intervention group were centrifuged twice at 2,500 g for 20 min. Large-sized vesicles (lEVs) were separated by differential centrifugation, and thirty-seven EV surface epitopes were studied using the MACSPlex Human Exosome Kit (Miltenyi) following the manufacturer’s instructions, as previously described^1,2^. In short, this assay employs phycoerythrin and fluorescein isothiocyanate-labelled polystyrene capture beads that capture plasma lEVs during overnight incubation. Subsequently, allophycocyanin-labelled anti-CD9, anti- CD63, and anti-CD81 antibodies were added to positively select for lEVs. This results in the formation of a structure encompassing capture beads, lEVs, and detection antibodies, facilitating event detection and identification of surface epitopes. Samples were measured using an Attune NxT flow cytometer, further analyzed using Flowjo (v10.10.0) and median fluorescence intensity (MFI) values were exported for statistical evaluation.

### Statistical analyses

Statistical analyses were performed using GraphPad Prism 10 (v10.1.1(270)). Data distribution was assessed with D’Agostino & Pearson and Shapiro-Wilk tests. Differences between two timepoints were assessed using the Wilcoxon matched pairs signed rank test. Mixed-effects analysis, with Geisser-Greenhouse correction and Tukey’s multiple comparison test was performed for the comparison of three time points. Sex-specific analyses were conducted to determine whether the changes observed in the overall population were also present when analyzed separately by sex. Graphical data representations were created using GraphPad Prism 9 and BioRender. An alpha value of *p* < 0.05 was used for all statistical tests. Statistical significance was set at *p* ≤ 0.05 and marked with asterisks as follows: * *p* ≤ 0.05, ** *p* ≤ 0.001, and *** *p* ≤ 0.0001. Data are presented as the arithmetic mean and corresponding standard error of the mean (SEM).

## Results

### Subproject 1

#### Acute moderate continuous exercise intervention affects monocyte activation 30 min post exercise in elderly individuals

To investigate the acute effect of moderate exercise (60 % VO_2max_ for 30 min), 14 participants were assessed before and 30 min after a moderate exercise intervention (Figure 1A-B). Flow cytometric analysis of whole blood revealed no significant alterations in cell composition or frequency of adaptive or innate immune populations following acute continuous moderate-intensity exercise (Figure 1C-D and Table 1). However, specific changes were observed in the monocyte subsets. Classical monocytes exhibited increased frequency of CD86 (pre, 20.12 ± 2.83 %; post, 26.97 ± 2.70 %, *p* = 0.0105) and decreased CX3CR1 expression post-intervention (pre, 54.27 ± 2.37 %; post, 46.81 ± 3.30 %, *p* = 0.0345). Intermediate monocytes displayed decreased CX3CR1 expression (pre, 80.12 ± 3.11 %; post, 73.40 ± 4.52 %, *p* = 0.0345) and reduced HLA-ABC levels (pre, 100 ± 0.11 %; post, 99.36 ± 0.19 %, *p* = 0.0219). No discernible changes in the nonclassical subset were observed (Figure 1E). Inflammatory cytokines showed no significant differences between baseline and post-intervention groups (Supplementary Figure 1).

**Table 1.** Whole blood events per microliter for every cell type studied before and after the exercise intervention. Values are presented in events/uL, showing mean and standard error of the mean (SEM) per cell group within each group. Statistical evaluation was performed using student’s t test, *p* values are shown within the table.

|  | baseline | 30 min |  |
| --- | --- | --- | --- |
| *n* | 14 | 14 | *t-test* |
|  | *mean of events per ul (SEM)* |  | *p-value* |
| *NKT cells* | 33.25 (4.5) | 34.03 (5.4) | 0.9598 |
| *NK cells* | 46.74 (5.2) | 53.4 (6.9) | 0.4879 |
| *B cells* | 27.2 (3.4) | 26.0 (3.0) | 0.8036 |
| *Cytotoxic T cells* | 12.1 (1.5) | 11.7 (5.3) | 0.9189 |
| *T helper cells* | 163.7 (23.8) | 145.9 (17.3) | 0.6027 |
| *Neutrophils* | 146.9 (15.4) | 146.6 (8.6) | 0.3380 |
| *Classical Monocytes* | 2.3 (0.2) | 2.14 (0.2) | 0.4542 |
| *Intermediate Monocytes* | 2.51 (0.3) | 2.59 (0.3) | 0.8300 |
| *Nonclassical Monocytes* | 5.71 (0.4) | 6.87 (0.7) | 0.3064 |

### Sex specific activation of monocyte subsets and IL-6 post exercise response

Due to the scarcity of sex-disaggregated data on exercise interventions in healthy elderly populations, we aimed to investigate potential sex-specific differences in the immune response. While overall immune cell counts between male and female participants did not differ significantly, when examining the activation of monocyte subsets, our results showed sex-specific variations following moderate exercise (Figure 7A). Male participants exhibited a notable decrease in CCR2 expression in classical monocytes and a significant increase in CXC3CR1 levels in intermediate monocytes. In contrast, female participants displayed reduced CD62L expression in intermediate monocytes. Additionally, only female participants exhibited an increase in circulating IL-6 levels post-exercise indicating a sex-specific inflammatory response to moderate exercise (Figure 7B).

### Subproject 2

#### Acute high-intensity exercise intervention affects monocyte, NK cell and neutrophil changes 24 h post exercise in elderly individuals

To evaluate the effects of an acute high intensity exercise session, 19 healthy participants, with a median age of 67 years and a near-equal distribution of sex (47.3% females), underwent a maximum-intensity cardiopulmonary exercise test (CPET). The acute high- intensive exercise intervention was designed to push participants to their physical limits, enabling the assessment of immune responses and extracellular vesicle release under maximal stress conditions (Figure 2A). Participants exhibited a mean BMI of 24.6 and a mean power output of 126 Watts (ranging from 50-250W) during the intervention, which displayed substantial inter-individual variability (Figure 2B). The exercise duration and VO_2max_ displayed notable similarities among participants, revealing the diverse fitness levels within the cohort (Figure 2C). All participants performed same CPET protocol. To evaluate the impact of intense exercise on immune cell populations, we conducted flow cytometric analysis was performed on blood samples collected at baseline, 30 minutes post-intervention and 24 h post-exercise (Figure 2A). This gating strategy allowed for detailed examination of neutrophils, T helper cells, cytotoxic T cells, NK cells, B cells, and monocyte subsets (Figure 3A). Our results revealed significant changes in the cellular fractions at the different time points, with the most prominent difference observed at 24 h post-intervention (Figure 3B). Specifically, our findings indicated a significant increase in classical and nonclassical monocytes, along with a rise in NK cells driven by CD56^bright^ and CD16^low^ NK cells contributing to the increase (Figure 3C). In contrast, neutrophil counts declined 24 h after the intervention, while components of the adaptive immunity remained unchanged (Figure 3C). Further sex-disaggregated analysis revealed that the increase in classical monocytes and NK cells was predominantly driven by the male participants, whereas no significant changes were observed in the female subgroup (Figure 7C).

**Figure 3.**
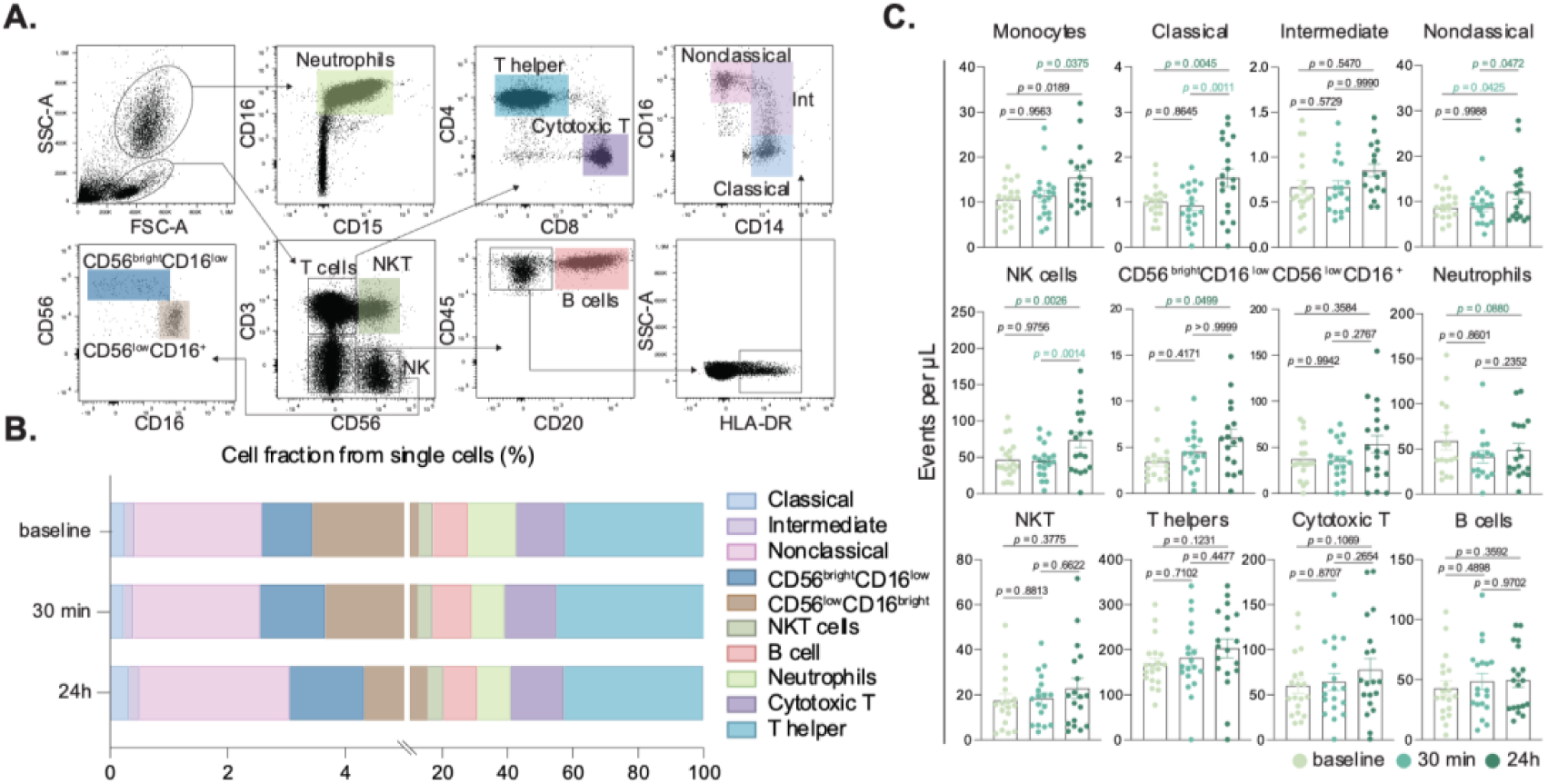
Peripheral immune cell subset alterations induced by intense exercise. Representative gating strategy to identify single cells, followed by granulocytes and mononuclear cells based on size and granularity (SSC-A vs FSC-A). Neutrophils were gated based on FSC and SSC, followed by their expression of CD16 and CD15. Mononuclear cells were gated on the expression of CD3 and CD56, to obtain NKT cells (CD56^+^, CD3^+^), NK cells (CD56^+^, CD3^-^). Further, NK cells were gated on the intensity of CD16 and CD56 expression, to subdivide them in CD56^bright^, CD16^low^ NK cells and CD56^low^, CD16^+^ NK cells. From the double negative population (CD56^-^, CD3^-^), we assessed the presence of CD20 to represent B cells, and the CD20-cells, were further gated based on their surface expression of CD16 and CD14, where classical monocytes were defined as CD14^+^, CD16^-^; intermediate monocytes were CD14^+^, CD16^low^ and, nonclassical monocytes as CD16^++^, CD14^low^ **(A)**. Bar charts showing cell fractions of all investigated immune cell populations at baseline (mint green), 30 min (turquoise) and 24 h (dark green) after the intervention **(C)**. Statistical tests were performed using mixed-effects analysis, with Geisser-Greenhouse correction and Tukey’s multiple comparison test. *p* values are shown in figure.

### Monocyte and neutrophil activation after acute intense exercise

Monocyte and neutrophil activation after acute intense exercise was investigated to determine if their activation status was influenced by the intervention. The expression levels of key activation markers including CCR2, CD36, CD62L, CD86, CD163, CX3CR1, and HLA-ABC were evaluated across each monocyte subset. In classical monocytes (Figure 4A), CX3CR1 expression was significantly reduced 30 min post intervention. In contrast, intermediate (Figure 4B) and nonclassical (Figure 4C) monocytes did not exhibit significant changes in the studied markers. For neutrophils, a significant decrease in CD62L expression was observed between the 30 min and 24 h time points (Figure 4D).

**Figure 4.**
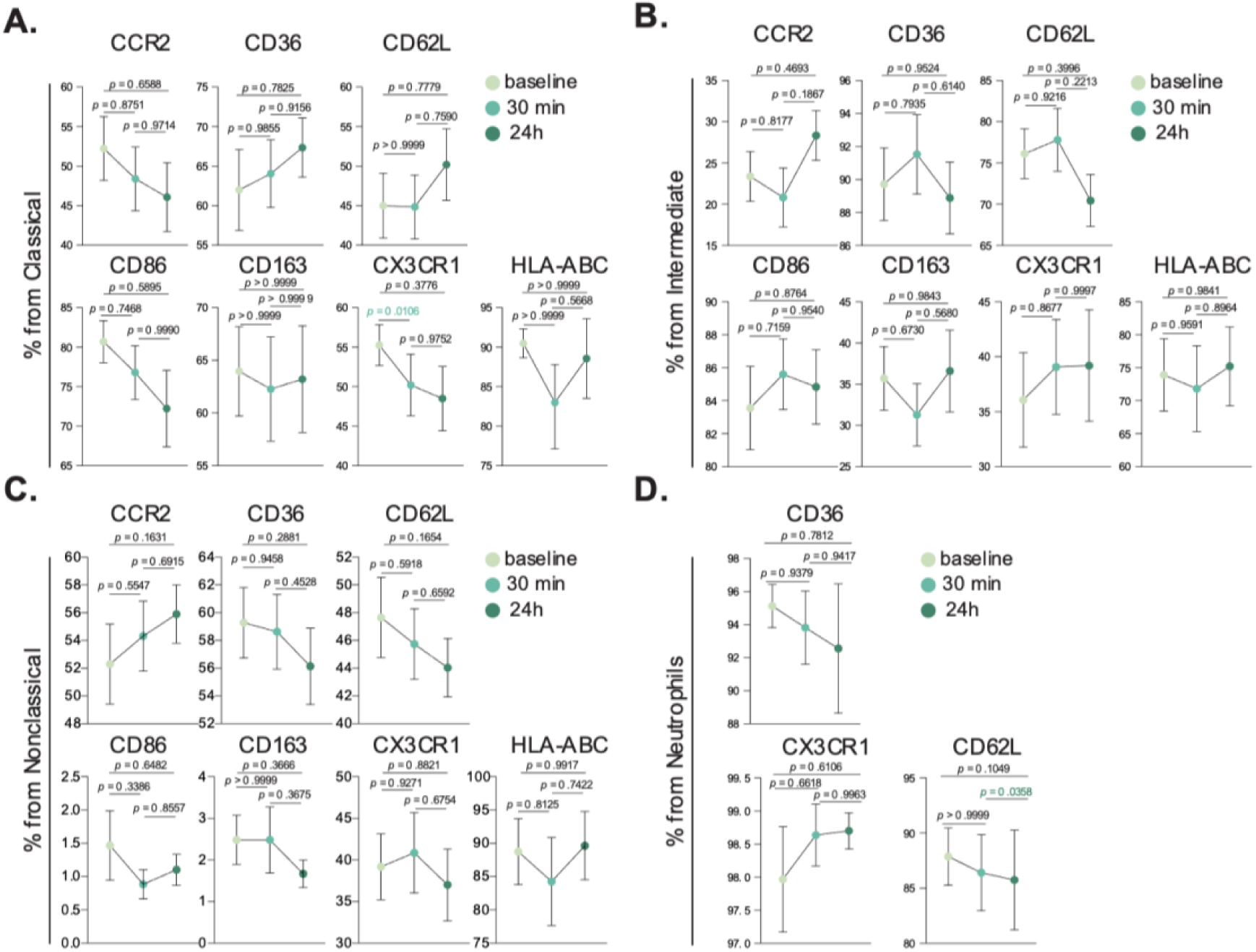
Activation dynamics of innate immune cell subsets following intense exercise. Bar charts showing cell fractions of all investigated activation markers in classical monocytes (A), intermediate monocytes (B), nonclassical monocytes (C) and, neutrophils (D), at baseline (mint green), 30 min (turquoise) and 24 h (dark green) after the intervention **(C)**. Statistical tests analyzed using a mixed-effects analysis, with Geisser-Greenhouse correction and Tukey’s multiple comparison test. *p* values are shown in figure.

### Elevated proinflammatory cytokine levels following acute high intensity exercise

Aging is linked to elevated levels of inflammatory cytokines, creating an environment conducive to age-related diseases^29^. Additionally, it has been shown that intense acute exercise in young individuals typically triggers a surge in canonical proinflammatory cytokines^30^, and we aimed to determine if similar responses occur in an elderly cohort. Therefore, to assess the inflammatory impact of high-intensity exercise, we measured various cytokines and regulatory biomolecules at baseline, 30 minutes and 24 hours post-intervention (Table 2). Our study revealed that high-intensity exercise significantly increased peripheral TNFα levels immediately post-exercise (5.32 pg/mL) compared to baseline (2.68 pg/mL, p = 0.0470), and remained elevated 24 h post-intervention (4.23 pg/mL), evidencing a sustained inflammatory response (Table 2). IL-6 levels did not show significant changes across time points, although a slight decrease was observed at 24 h post-intervention. Other cytokines and biomarkers, including BDNF, IL-23, IL-1β, and IL-12p70, did not show significant fluctuations post-exercise. Similarly, markers associated with vascular and brain health, such as sRAGE, sTREM2, and β-NGF, did not exhibit significant changes. However, a separate analysis showed a significant correlation of BDNF levels with age 24 h after the continuous acute intense exercise intervention (Figure 5). Anti-inflammatory cytokine IL-10 showed a transient non-significant decrease immediately post-intervention returning to baseline levels by 24 h (Table 2).

**Figure 5.**
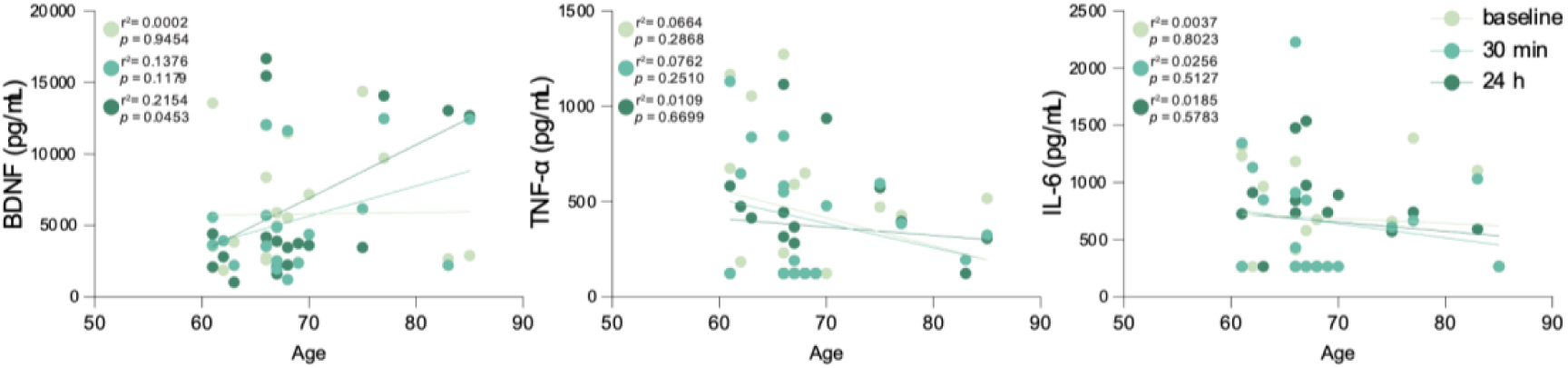
Peripheral BDNF levels positively correlate with age in elderly participants 24h following acute continuous intensive exercise. Scatter plots displaying peripheral levels of brain-derived neurotrophic factor (BDNF), tumor necrosis factor-alpha (TNF-α), and interleukin-6 (IL-6) in relation to age across the three studied time points: baseline (mint green), 30 min post-exercise (turquoise), and 24 h post-exercise (dark green). Data points represent individual participants, with green hues indicating different time points as specified. r^2^ and *p* values are shown in plot; statistical analysis was performed using simple linear regression.

**Table 2.**
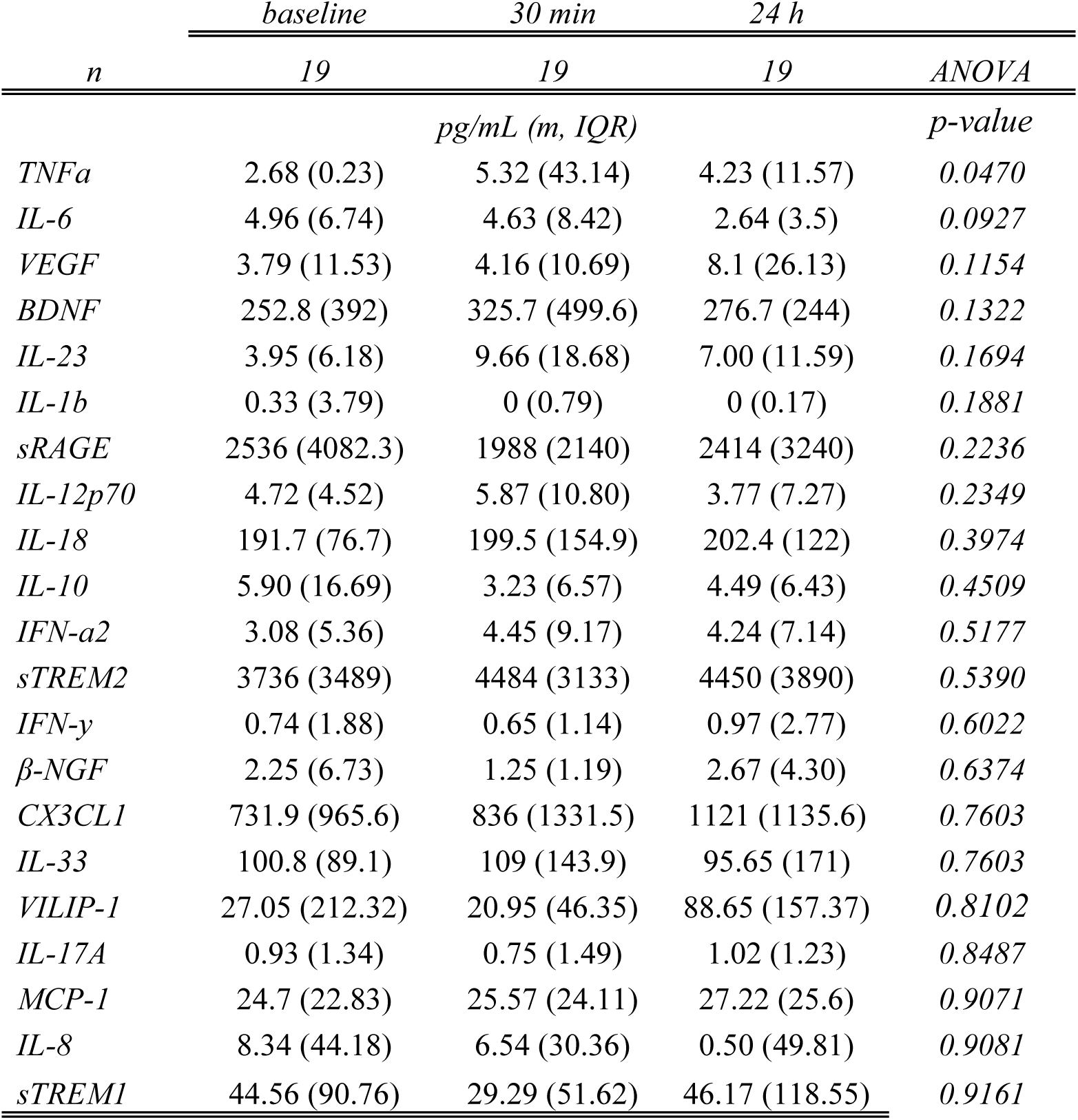
Circulating cytokine levels in the intense exercise intervention. Values are presented in pg/mL, showing median and inter-quartile range (IQR) per analyte within each group. Statistical evaluation was performed using mixed-effects analysis, with Geisser-Greenhouse correction and Tukey’s multiple comparison test, was performed for the comparison of baseline, 30 min, and 24 h after the intense exercise intervention.

### Increased extracellular vesicle release is a key response to intense exercise

Following the evaluation of immune cell populations, their activation status, and peripheral cytokine levels, we examined the impact of high-intensity exercise on the release of EVs. Plasma-derived lEVs were quantified at baseline, 30 minutes and 24 h post-intervention. Flow cytometric analysis demonstrated a general increase in the concentration of plasma-derived EVs after 24 hours, as indicated by increased levels of tetraspanin markers CD9, CD63, and CD81 (Figure 6A-B). Notably, CD63 and CD81 displayed significant increases in median fluorescence intensity (MFI) at 24 h compared to 30 minutes post-intervention, while CD9 levels remained unchanged.

**Figure 6.**
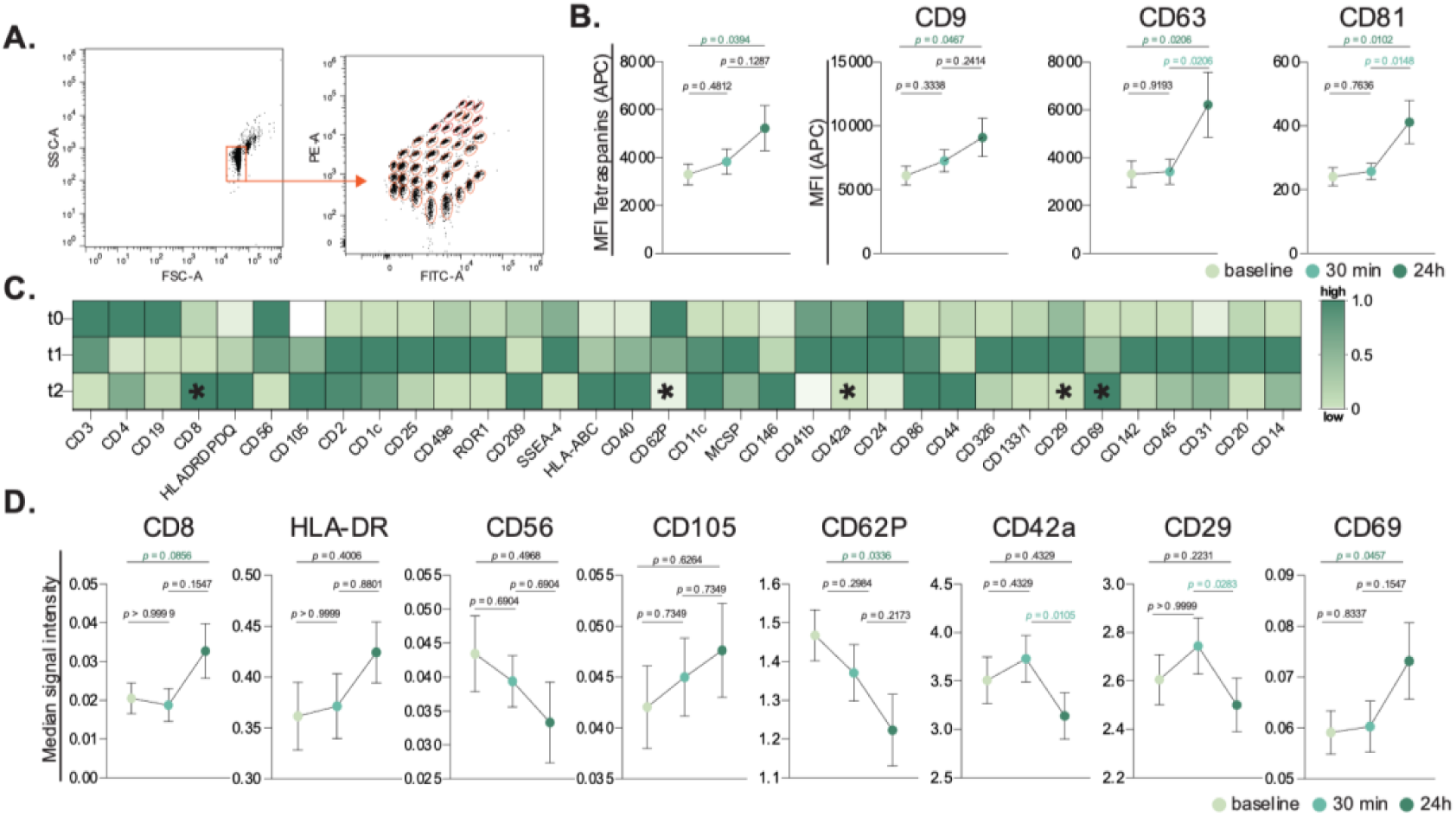
Extracellular vesicles release in response to acute continuous intensive exercise. Representative gating strategy of the bead-based extracellular vesicle assay, showing the bead size selection (left) and the 37 different dye-labeled capture beads against 37 different EV surface antigens based on FITC and PE fluorescence (right) **(A)**. Column plot showing mean and SEM of the tetraspanin signal (CD9, CD63 and CD81) (left) and individualized representation of CD9, CD63 and CD81 in each timepoint studied (right) **(B)**. Heatmap showing CD9-CD63-CD81 normalized MFI in all individual samples, showing 34 differentially expressed surface markers in our cohort before, 30 min and 24 h after the intervention **(C)**. Column plots showing mean and SEM of signal intensity of lymphocytes and leukocytes (CD8, HLA, CD56), endothelial cells (CD105), platelets (CD62P, CD42a) and, immune cell activation (CD29, CD69) after detection of CD9, CD63 and CD81. Statistical tests analyzed using a mixed-effects analysis, with Geisser-Greenhouse correction and Tukey’s multiple comparison test **(D)**. *p* values are shown in figure.

**Figure 7.**
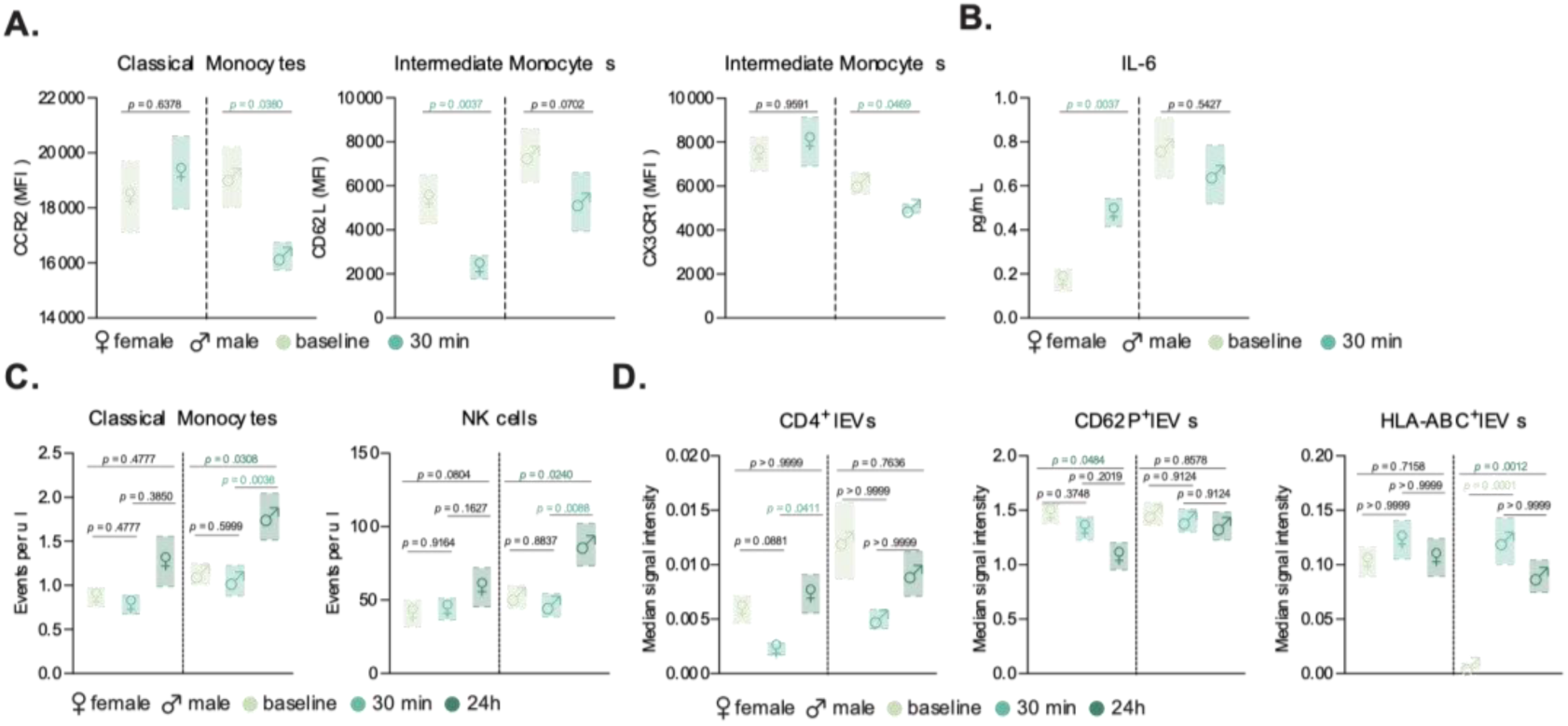
Sex-specific immune and EV responses to exercise in older adults. Floating bar charts showing sex-disaggregated (female symbol on the left and male symbol on the right) mean and SEM of activation markers in monocyte subsets after a moderate exercise intervention **(A)** and circulating IL-6 levels in pg/mL **(B)** comparing baseline and 30 min after the intervention. Flowing bar chart showing sex-disaggregated data (female symbol on the left and male symbol on the right) mean and SEM of events per microliter of classical monocytes (left) and NK cells (right) **(C)**, and median signal intensity of CD4^+^, CD62P^+^ and HLA-ABC^+^ ExerVs **(D)** at baseline, 30 min and 24 h after the intense exercise intervention. Statistical evaluations for the moderate intervention comparing two timepoints was performed using non-parametric student’s t-test (n=14), and for the intense exercise intervention data, comparing three timepoints a mixed-effects analysis, with Geisser-Greenhouse correction and Tukey’s multiple comparison test was performed (n=19). *p* values are shown in figure.

A detailed examination of 37 exosomal surface epitopes revealed a significant increase in surface CD69 expression after 24 h post-intervention (Figure 6C). This increase in CD69, a marker of cellular activation, was accompanied by a significant reduction in CD62P, CD42a, and CD29 markers on lEVs at 24 h post-intervention. Other markers including CD8, CD56, and CD105, did not show significant changes across the time points (Figure 6D). Sex-specific differences were also observed in lEV profiles. While the total circulating lEV count did not differ between sexes, females exhibited a significant increase in CD4+ lEVs and a decrease in CD62P+ lEVs 24 hours post-exercise. In contrast, male participants showed a marked increase in HLA-ABC+ lEVs following the intense exercise intervention (Figure 6D).

## Discussion

In recent years, the *Exercise as Medicine* approach has been increasingly recognized for its multifaceted benefits beyond disease prevention. Exercise acts as a modulator of immune function and inflammation, key elements in aging and disease processes^27,31,32^. Therefore, our study aimed to explore the interplay between exercise and the immune response, revealing alterations in immune cell populations and cytokine levels following both acute continuous moderate and intense exercise interventions in elderly individuals. Furthermore, we examined exercise induced release and selective surface marker modulation of ExerVs as potential mediators of systemic immune response, contributing to a growing body of evidence that exercise plays a vital role in promoting healthy aging and preventing age-related diseases.

### Monocyte subsets activation after moderate exercise

In the initial phase of our study, we sought to investigate the impact of a single session of acute continuous moderate exercise on the peripheral innate immune system of healthy elderly individuals. While no significant changes were detected in peripheral cell numbers or distribution, distinct alterations emerged in the activation of monocyte subsets. Previous studies indicate that monocyte subset activation in response to exercise depends on the exercise intensity^33^. Our findings align with this, as classical (CD14^+^CD16^-^) monocytes, representing an early differentiation stage are strongly recruited into circulation during long-term aerobic exercise (in our study acute continuous moderate exercise: 30 minutes, 60 % VO_2max_, subproject 1). In contrast, mature subtypes (non-classical and intermediate monocytes, CD14^+^CD16^+^) increase with high intensity exercise above the individual anaerobic threshold (subproject 2). Given that aging is associated with chronic innate immune activation – particularly a higher frequency of CD16^+^ monocyte subpopulations – exercise emerges as a primary intervention target for healthy aging^34^.

Classical monocytes, which are primarily responsible for phagocytosis and the initiation and mediation of leukocyte recruitment, displayed an increase in CD86 and a decrease in CX3CR1 expression. CD86 is a co-stimulatory molecule that is rapidly upregulated upon stimulation, as it is present in a reservoir in monocytes and dendritic cells, which in this context allows for quick deployment to the cell surface for enhanced co-stimulatory function^35^. Chemokine receptor CX3CR1, which binds to CX3CL1, has been demonstrated to play a role in regulating monocyte differentiation and migration. Research has revealed that CX3CL1 levels are elevated in human skeletal muscle following physical activity, which is believed to facilitate the creation of a regenerative microenvironment post-exercise^36^. Intermediate monocytes also exhibited decreased CX3CR1 levels post-exercise. This decrease has been associated with the differentiation process that monocytes can go through in the periphery, which is also evidenced in our cohort by increasing levels of intermediate and nonclassical monocytes compared to classical monocytes levels after the intervention^37^. Furthermore, our study documented a significant reduction in the surface expression of monocyte chemokine receptor 2 (CCR2) among classical monocytes in male participants following exercise. CCR2, a chemokine receptor critical for monocyte recruitment, has shown sex-specific expression patters in previous studies with younger females exhibiting higher expression levels compared to males after a single instance of physical activity^38,39^. Consistent with this finding, our study revealed higher baseline CCR2 levels in elderly females compared to their male counterparts. Specifically, our results evidenced a considerable reduction in CCR2 expression following exercise exclusively in elderly males. This sex-specific modulation of CCR2 accentuates inherent biological differences, potentially influenced by hormonal factors, significantly modulating the immune system responses to exercise between elderly men and women. This finding presents the potential value for personalized regimens of exercise to optimize immune health in the aging population.

Our analysis of intermediate monocytes revealed a concomitant decrease in the expression levels of CX3CR1 and HLA-ABC following moderate exercise. HLA-ABC, also known as the major histocompatibility complex class I antigen (MHC-I), plays a crucial role in antigen presentation and has been demonstrated to increase its microglial expression with age, suggesting that the immune response in the elderly may be subject to modification, consequently increasing their susceptibility to structural brain changes^40,41^. However, in the peripheral monocyte subpopulation, the observed decrease in HLA- ABC levels after moderate exercise in the elderly suggests a possible transient period of heightened susceptibility to infections. Moreover, our results indicated a notable reduction in the expression of CD62L, an adhesion molecule critical in the recruitment of monocytes to sites of inflammation through endothelial adhesion and subsequent tissue migration^42,43^. Prior studies have reported higher expression of CD62L on peripheral blood mononuclear cells (PBMCs) in older individuals as compared to younger cohorts ^44^. Interestingly, in our study, this reduction in CD62L levels was observed only in female participants following moderate exercise. This finding suggests shedding of CD62L from the monocyte surface as a consequence of a more efficient monocyte adhesion and transmigration to sites of inflammation potentially enhancing their acute inflammatory response in the surrounding tissues. In contrast, the relatively stable CD62L levels observed in male participants suggest that their monocytes may be less efficient in transitioning from rolling to firm adhesion and transmigration under acute inflammatory conditions. This may contribute to a higher chronic inflammatory state, potentially explaining the higher incidence of cardiovascular disease observed in men compared to women.

### Circulating pro-inflammatory cytokines after one moderate session of exercise in the elderly

Prior research demonstrates that acute engagement in moderate-intensity physical activity can result in an increase in proinflammatory cytokines, including interleukin-6 (IL-6) and tumor necrosis factor-alpha (TNF-α) ^30,45,46^. Contrary to these studies, we did not observe significant increases in proinflammatory cytokines immediately after post-moderate exercise, possibly due to the participants’ familiarity with moderate activity or the timing of the assessment. According to the Borg RPE Scale, the participants reported an average score of 15 (ranging from 13 to 18), indicating that the perceived exertion level ranged from somewhat hard to very hard, which should have been sufficient, theoretically, to induce a transient proinflammatory state^47^. We hypothesize that this finding could be explained by the fact that the assessment was conducted 30 minutes after the intervention, which did not provide sufficient time for the signaling, production, and release of proinflammatory markers. However, this could also be due to the participants’ tolerance to moderate exercise, as most of them reported engaging in regular physical activity and were not sedentary individuals.

We detected elevated IL-6 exclusively in the female cohort. Previous research indicates that younger and middle-aged women with ischemic heart disease tend to exhibit higher IL-6 concentrations than men with the same condition, particularly in response to stress, a difference potentially influenced by testosterone^48^. In aging populations, plasma IL-6 levels are negatively correlated with muscle strength in both men and women. However, men with better muscle condition show higher levels of IL-6 than women with poorer muscle condition, indicating a sex-specific disparity in IL-6 as a predictive marker for sarcopenia^49^. Our findings revealed that moderate aerobic exercise led to a substantial increase in plasma IL-6 among females, suggesting that this exercise intervention may elicit different inflammatory responses in elderly women versus men. This sex-specific IL-6 response could have important implications for inflammation regulation and health outcomes in aging populations.

### Intense exercise modifies the innate immune response in the aging population in comparison to acute moderate continuous exercise

Exercise-induced leukocyte mobilization is well documented, and it is postulated that cortisol release during the intervention facilitates the exit of leukocytes from the bone marrow^50,51^. We hypothesized that a single session of intense exercise would provoke a more pronounced effect in the peripheral immune system compared to moderate exercise. Early studies indicate that NK cells are the first and most affected immune cell subset after exercise^30,52–55^. Consistent with these studies, we confirmed an enhanced innate immune response, particularly in the levels of NK cells and monocytes 24 hours after the intervention. Furthermore, we conducted a study analyzing the expression of CD56 and CD16 in mature natural killer (NK) cells, specifically focusing on the CD56^bright^ and CD16^low^ subsets, which were found to be the main contributors to the increased NK cell count 24 hours after exercise. Unlike moderate exercise, which did not alter proinflammatory cytokine levels in the elderly population, intense exercise led to a significant increase in TNFα. This rise in TNFα is likely attributed to a specific subset of NK cells, known for their elevated proinflammatory cytokine production which aligns with our data on heightened TNFα levels sustained for up to 24 hours post- intervention^56,57^. These data indicate that intense exercise triggers a pronounced acute inflammatory response in aging individuals, particularly through elevated TNFα levels, while other cytokines display trends that suggest additional underlying immunological effects. Further studies with larger cohorts are needed to fully elucidate the broader immunological impact of intense physical activity in elderly individuals.

Our results demonstrate a significant increase in the levels of classical and nonclassical monocytes 24 h post-intense exercise. Analysis of activation markers revealed a decline in CX3CR1 expression on classical monocytes, mirroring the decreased observed after moderate exercise. CX3CR1 decrease in this context could hint towards a shift towards a more proinflammatory phenotype in response to intense exercise. As expected, there was a reduction in the number of neutrophils observed at the 24-hour timepoint. This decrease is thought to be a result of neutrophils being recruited to muscle tissue in response to exercise-induced tissue damage^58,59^. This aligns with previous findings by Nunes-Silva et al. who reported that exhaustive treadmill exercise in mice induced rolling, adhesion, and transmigration of neutrophils into muscle tissue^60^. Consistently, our cohort showed increased expression of CX3CR1 in neutrophils a mechanism that has been shown in mice to facilitate neutrophil recruitment to active muscles during exercise^61^.

Research has consistently shown that physical exercise has a significant impact on BDNF levels. Both acute (single session) and regular (long-term) exercise have been demonstrated to increase BDNF levels, which is associated with cognitive improvements^62^. Furthermore, it has been shown, that acute exercise modulates BDNF and pro-BDNF Protein content in immune cells^63^. Aerobic exercise has been shown to increase peripheral BDNF levels in elderly adults^64^, and in our cohort, BDNF levels exhibited a slight increase 30 min after the intense exercise intervention, with levels nearly returning to baseline by the 24 h timepoint, which is consistent with prior findings. Notably, when we examined the correlation between BDNF levels and age, we observed no significant correlation at baseline or 30 min post-intervention. However, a significant positive correlation emerged at the 24 h timepoint. This finding indicate that the sustained elevation of BDNF levels 24 h after intense exercise is more pronounced with increasing age, suggesting an age-dependent response in BDNF dynamics post-exercise. This could be driven by age-related differences in the sensitivity of BDNF production and release in response to exercise stress over time. Alternatively, older individuals might exhibit prolonged BDNF elevation as a compensatory neuroprotective mechanism in response to the demands of intense exercise. Whether the delayed BDNF response correlates with improved recovery or neuroplasticity in older individuals needs to be further explored.

### Extracellular vesicle release after exercise in the elderly

EVs are crucial in mediating the beneficial effects of physical activity by transporting proteins, lipids, metabolites, and nucleic acids into the circulation. These vesicles facilitate communication with recipient cells and modulate intercellular communication and metabolic processes^65–68^. In 2019, Brahmer et al. investigated the release and origin of circulating EVs following exercise in a cohort of healthy male athletes. Their findings revealed a significant increase in the number of EVs released during exercise, primarily attributed to lymphocytes, monocytes, platelets, endothelial cells, and MHC-II+ cells ^69^.

In our elderly cohort, both males and females exhibited an increased release of ExerVs from lymphocytes and leukocytes (CD8, CD14, HLA, CD86), platelets (CD62P, CD42a, CD41b), and endothelial cells (CD105, CD31). This finding aligns with previous research conducted exclusively on younger, healthy participants, emphasizing the relevance of understanding ExerV dynamics in older adults and their potential implications for immune function and health in aging populations^69,70^. Furthermore, we observed a significant increase in CD69 expression 24 hours post-exercise. CD69 expression is upregulated upon activation in leukocytes, including NK cells and monocytes, which coincide with their increased numbers found in circulation ^71^. Additionally, an increase in platelet marker expression (CD62P, CD42a) on ExerVs was observed 30 min after exercise, but showed a significant reduction at the 24-hour time point. This observation suggests that exercise induces acute platelet activation leading to the release of ExerVs and could indicate that the transient increase in platelet-derived ExerVs may contribute to the exercise adaptive response and support recovery processes post-exercise in aging individuals.

While no differences were observed in the total amount of ExerVs release between females and males in our cohort, sex-specific analyses revealed distinct patterns. In females, a significant increase in T cell-derived ExerVs (CD4+) was detected, whereas males exhibited a significant increase in MHC-I ExerVs (HLA-ABC+) 24 hours after the exercise intervention. These findings suggest adaptive immunity alterations that had gone unnoticed at a cellular level potentially influenced by hormonal factors. Estrogen for example, could potentially enhance CD4+ T cell activation leading to higher levels of CD4+ EVs in females, while testosterone may promote antigen presentation and cytotoxic responses^72^, as reflected by the increase of HLA-ABC+ ExerVs in males. Additionally, sex-specific physiological adaptations to exercise could contribute, with females exhibiting a stronger immune modulation response and males focusing on recovery and inflammation resolution. Nevertheless, whether these differences stem from hormone-dependent effects, muscle fitness, or metabolic mechanisms, remains to be determined. Our findings indicate that adaptive immune responses to exercise-induced ExerVs release may vary in a sex-dependent matter, warranting further research to explore those mechanisms.

## Conclusion

Physical exercise impacts several physiological systems beyond solely skeletal muscle engagement^9,73^. Due to its pivotal role in human health and disease management, exercise has garnered substantial attention for its numerous beneficial effects, particularly its role in promoting healthy aging. As the global population ages, addressing the challenges of healthy aging becomes crucial not only from an individual health perspective but also from economic and societal standpoints^74^. Regular physical activity has been demonstrated to delay the onset of age-related disorders such as cardiovascular diseases, atherosclerosis, arterial hypertension and dementia^75–78^. Since physical inactivity correlates with an increase in health care costs, regular exercise is a cost-effective method to reduce the financial and resource burden on the health care system and improve the individual’s quality of life^79^.

In summary, our study further defines the complex interplay between exercise and immune function in elderly individuals, demonstrating significant alterations in immune cell populations, cytokine production, and EV release in response to both moderate and intense exercise. These findings point towards the necessity for tailored exercise prescriptions that consider individual physiological responses, optimizing health outcomes within aging populations. Notably, the increase concentration of plasma-derived EVs and variations in surface marker expression suggest a multifaceted signaling network that influences innate immune modulation, platelet function, and the body’s adaptability to physical activity. Our results position EVs not only as biomarkers of exercise-induced physiological changes but also as potential targets for therapeutic interventions. As the *Exercise as Medicine* paradigm continues to evolve, our research advocates for a deeper understanding of exercise’s role in enhancing immune function and resilience against age-related inflammation, paving the way for innovative health strategies in the aging population. The data indicate that both moderate and intense exercise induce significant changes in immune cell activation and cytokine levels in this healthy elderly population. Given the compelling data supporting the multifaceted benefits of regular physical activity, we recommend the integration of exercise into daily routines, especially for the elderly. Considering sex- specific physiological differences when tailoring exercise regimens may further enhance health outcomes and optimize immune responses. Future research should continue to explore the process of exercise-induced immune modulation and explore the potential therapeutic applications of exercise in managing aging-related health challenges.

## Data Availability

All data produced in the present study are available upon reasonable request to the authors

## Table of contents category

*Exercise*

## Author Contribution

APG drafted the manuscript, conducted all experiments and analyses, and created the figures. APG and LM interpreted and visualized the data. ALM performed and helped analyze experiments. MS and YL recruited participants and conducted the exercise interventions. SS, CP, RBD, and LM contributed to manuscript preparation. PM designed the study, supervised the intervention, project administration, and revised the manuscript. IRD supervised the experiments, project administration, and the manuscript preparation. Each author is accountable for the work and for addressing any questions related to accuracy or integrity. All authors have reviewed and approved the final submitted version, meet authorship criteria, and contributed to manuscript revisions.

## Ethical approval

The study protocol was approved by the Ethics Committee of the Medical Faculty at Otto-von-Guericke University in Magdeburg, Germany (reference number: 07/20). All the procedures were conducted in accordance with the ethical standards outlined in the Declaration of Helsinki. Prior to participation, each individual provided written informed consent following a comprehensive explanation of the study, and all queries were addressed satisfactorily.

## Supplementary data

**Supplementary Figure 1.**
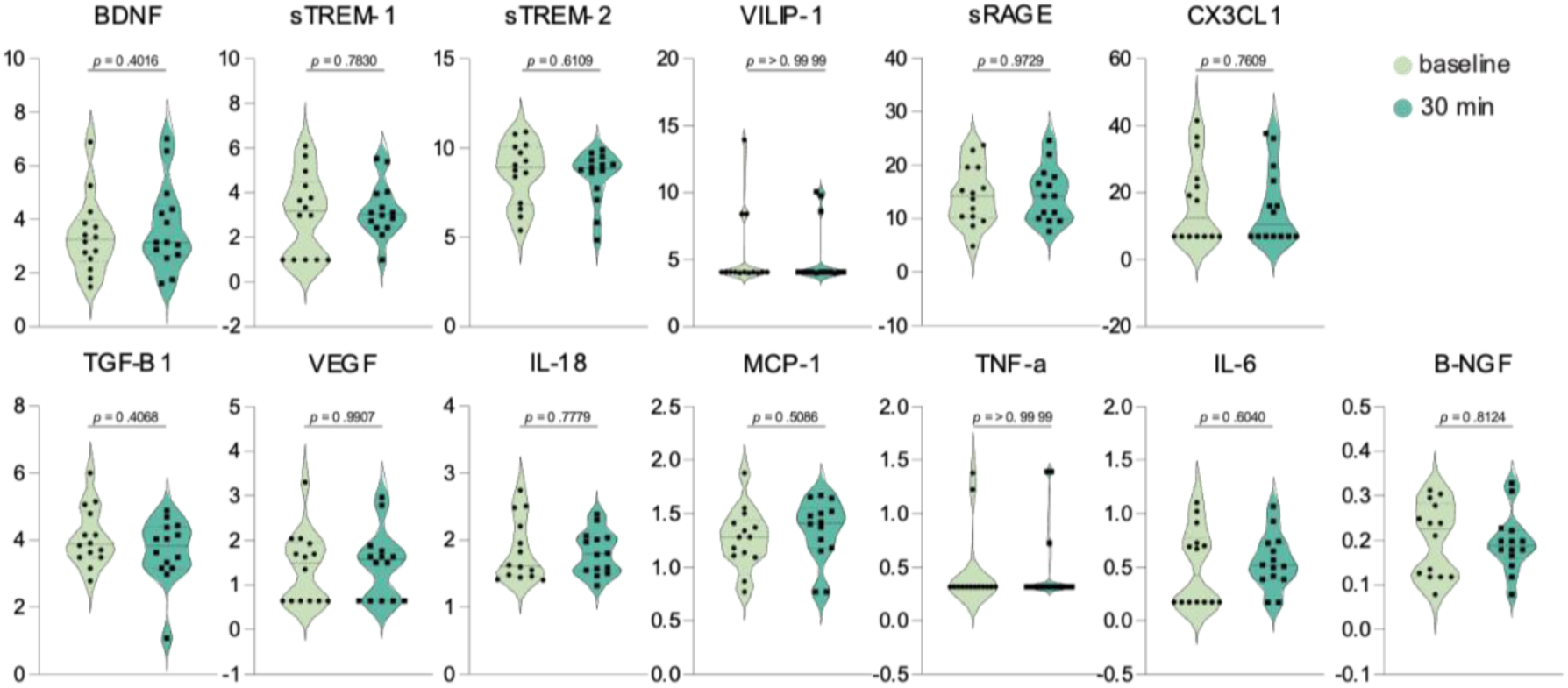
Peripheral cytokine levels 30 minutes following acute continuous moderate exercise. Violin plots display plasma concentrations of selected cytokines in elderly adults, measured at baseline and 30 minutes after a session of moderate-intensity exercise. Plots showing the central line representing the median value, and thinner lines above and below the median mark the interquartile range (IQR). Statistical evaluation was performed using student’s t test, *p* values are shown in figure.

